# Human germline biallelic loss-of-function *OSMR* variants cause severe allergic disease

**DOI:** 10.1101/2025.08.05.25332527

**Authors:** Mehul Sharma, Simran Samra, Yihui Liu, Alyssa James, Christina Michalski, Pariya Yousefi, Kate L. Del Bel, Henry Y. Lu, Ashish A. Sharma, Maja Tarailo-Graovac, Joshua Dalmann, Lily Buder, Bhavi Modi, Britt Drogemoller, Géraldine Blanchard Rohner, Christof Senger, Wingfield Rehmus, Julie S. Prendiville, Massimo Mangino, Colin J. Ross, Clara DM. van Karnebeek, Wyeth W. Wasserman, Pascal M. Lavoie, P M Prathibha, Catherine M. Biggs, Michael Boehnke, Leena Kinnunen, Heikki A. Koistinen, Margaret L. McKinnon, Siddaramappa Jagdish Patil, Diana K. Bayer, Jonathan J. Lyons, Stuart E. Turvey

## Abstract

OSMRβ (Oncostatin M receptor beta), a member of the IL-6 superfamily of cell surface receptors, binds OSM and IL-31 and plays a critical role in human immunity. We identified probands from four kindreds with biallelic damaging variants in *OSMR*, which encodes OSMRβ. Patients had a unifying phenotype for severe widespread, early-onset atopic dermatitis, peripheral eosinophilia, and elevated serum IgE. Patient OSMRβ variants were not appropriately expressed on the cell surface compared to OSMRβ^WT^. Patient *OSMR* variants showed significantly reduced OSM-mediated activation of STAT1, STAT3, and STAT5 and distinct transcriptional changes in primary dermal fibroblasts, including loss of interferon and inflammatory signatures. These defects were rescued upon lentiviral transduction of WT-*OSMR*. Together, these data establish that human germline biallelic loss-of-function *OSMR* variants cause severe allergic disease. We anticipate that this discovery will facilitate the recognition of additional affected individuals and the full definition of this novel primary atopic disorder.

## INTRODUCTION

Inborn errors of immunity (IEIs) are a group of single gene defects in which parts of the human immune system are missing or dysfunctional (Bousfiha et al., 2025). Primary atopic disorders (PADs) are a subset of the IEIs where severe allergic disease is the predominant clinical feature (Lyons and Milner, 2018). The discovery of new PADs advances our understanding of the molecular mechanisms responsible for allergic inflammation and can transform the clinical care of affected individuals (Chen et al., 2021; Lyons and Milner, 2018; Vaseghi-Shanjani et al., 2025).

In this study, we describe a novel human PAD caused by germline biallelic loss-of-function (LOF) variants in the gene *OSMR* found in 4 patients from 4 unrelated families spanning three continents. OSMRβ (Oncostatin M receptor beta), encoded by *OSMR*, is a member of the IL-6 superfamily. OSMRβ forms a heterodimeric component of both the OSM type II receptor (OSMRβ/GP130) and the IL-31 receptor (OSMRβ/IL-31Rα) and thus is integral in mediating the biological functions of oncostatin-M (OSM) and IL-31. OSM is a monomeric glycoprotein produced by activated T cells, monocytes (Zarling et al., 1986), macrophages (Repovic and Benveniste, 2002), and neutrophils (Cross et al., 2004; Goren et al., 2006), whereas IL-31 is mainly produced by CD4^+^ T helper cells (Dillon et al., 2004). Human biallelic OSM deficiency causes a bone marrow failure syndrome (Alfalah et al., 2025; Garrigue et al., 2025). The expression of the IL-31 receptor is specifically high in the dorsal root ganglion and is recognized for promoting itch in prurigo nodularis (Kwatra et al., 2023) and other pruritic skin conditions, while OSMRβ is widely expressed in a variety of cell types and tissues, including epithelial cells, fibroblasts, blood vessels, nerve cells, respiratory tissues, adipose tissues, and lymph nodes (Sonkoly et al., 2006) (Mosley et al., 1996). OSMRβ also plays a role in keratinocyte proliferation, differentiation, and inflammatory responsiveness (Heinrich et al., 2003). As such, signaling through the OSMRβ axis is important for various disease states (West, Owens, and Hegazy, 2018), including inflammatory bowel disease (Bachetti et al., 2021; Kim, Kaser, and Blumberg, 2017; West et al., 2017), pulmonary fibrosis (Mozaffarian et al., 2008), inflammatory skin conditions such as IL-31-mediated pruritus (Datsi et al., 2021), diffuse cutaneous systemic sclerosis (Stifano et al., 2018), cutaneous T cell lymphoma (Nattkemper et al., 2016), chronic autoimmune urticaria (Luo et al., 2018), and familial primary localized cutaneous amyloidosis (FPLCA) (Arita et al., 2008; Lin et al., 2010; Sakuma et al., 2009; Tanaka et al., 2010; Ueo et al., 2016; Wali et al., 2015). FPLCA is an extremely pruritic skin disorder characterized by amyloid deposits in the superficial dermis. Variants in *IL31RA* and *OSMR* are associated with FPLCA, which is typically an autosomal dominant disorder with onset of symptoms in adolescence or adulthood (Arita et al., 2008; Lin et al., 2010; Sakuma et al., 2009; Tanaka et al., 2010; Ueo et al., 2016; Wali et al., 2015).

Here we expand our understanding of the role of OSMRβ in human health and disease through this first description of patients with biallelic LOF variants in *OSMR* causing a new primary atopic disorder.

## RESULTS

### Identification of patients with biallelic *OSMR* deficiency and severe early-onset atopic dermatitis

We investigated four patients from four kindreds with severe early-onset atopic disease, most notably challenging to treat atopic dermatitis. Patients were identified by their clinicians as candidates for assessment of a monogenic disorder based on their severe phenotype (as detailed in the next section). All patients carried biallelic variants in *OSMR* (NM_003999.3). By sequencing their healthy parents and siblings (when available), we established an autosomal recessive pattern of inheritance. Readers can contact the corresponding author to request access to patient pedigrees.

Patient 1 (P1) (from Kindred A) carried compound heterozygous missense *OSMR* variants (c.1046C>A, p.A349D and c.1307T>A, p.V436D), which they inherited from their healthy heterozygous parents. Patients 2 and 3 (P2 and P3) (from Kindreds B and C, respectively) were homozygous for the c.1307T>A, p.V436D *OSMR* variant. P2 inherited the variants from their healthy heterozygous parents, while no pedigree information was available for P3. Patient 4 (P4) (from Kindred D) was homozygous for the c.1979_1980delAC, p.Y660Sfs*16 *OSMR* variant. P4 inherited the variant, which creates a premature stop codon, from their healthy heterozygous consanguineous parents.

All identified variants were located in the extracellular fibronectin III domains of the OSMRβ receptor, a region known to be important for receptor dimerization leading to signal transduction (Kurth et al., 2000; Timmermann et al., 2002) (**Fig. 1A**). Both the p.A349D and p.V436D variants result in the substitution of a nonpolar amino acid (alanine and valine, respectively) with a negatively charged amino acid (aspartic acid) at positions that are well conserved throughout evolution (**Fig. 1A**). The p.Y660Sfs*16 variant causes a premature stop codon. The p.A349D and p.Y660Sfs*16 variants have not been reported in any population databases or the Catalogue of Somatic Mutations in Cancer (COSMIC) database (**Fig. 1B**). The p.V436D variant has been reported in the genome aggregation database (gnomAD v4.1.0, non-UKB) with a monoallelic frequency of 2.47×10^-3^ and 5 homozygotes. All the *OSMR* variants were predicted to be damaging by a variety of *in silico* tools (**Fig. 1B**).

**Figure 1.**
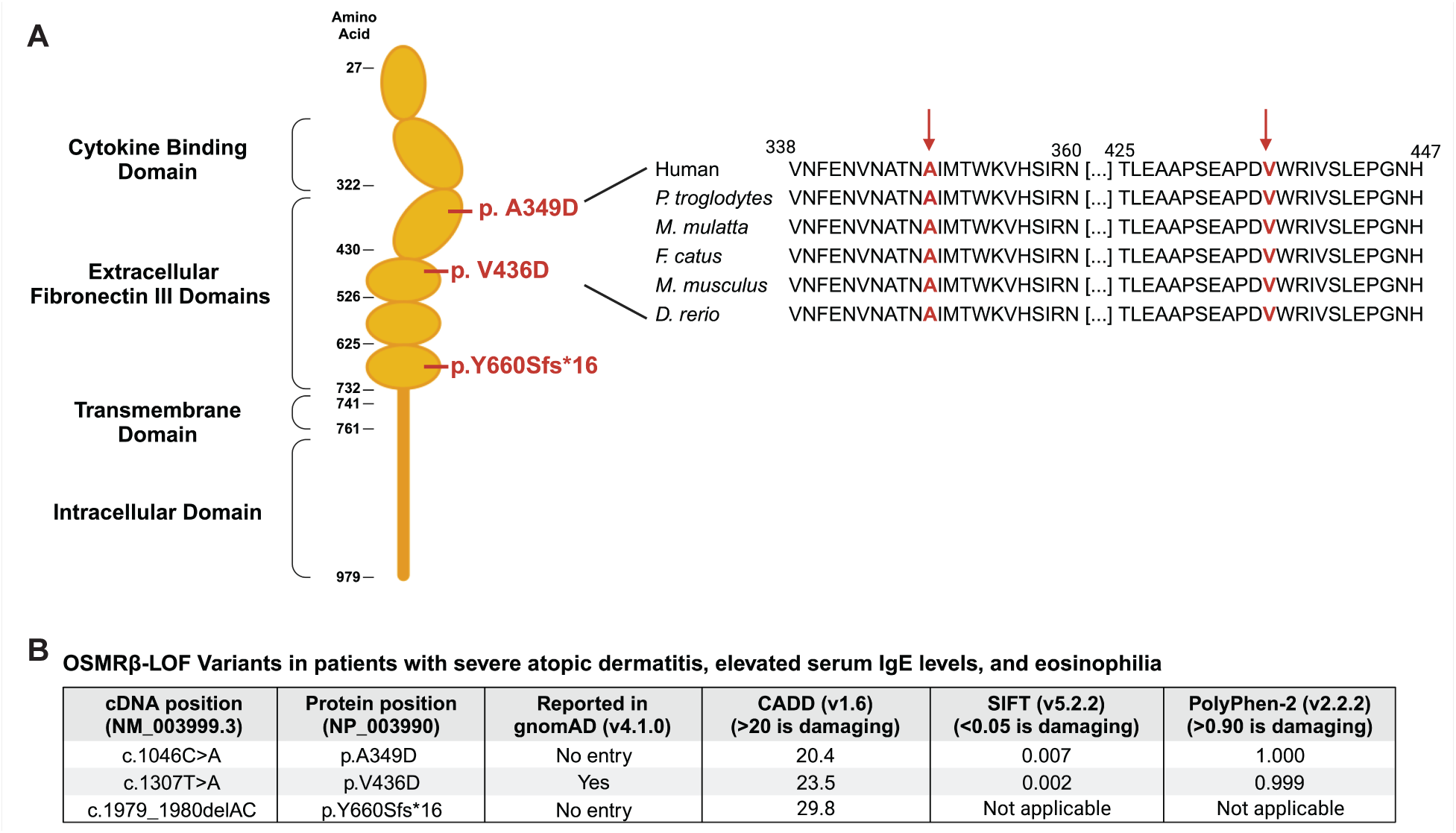
Four patients with severe allergic disease and *OSMR* variants. **(A)** Schematic illustrating the protein domains of OSMRβ. Amino acid location of the variants are shown in red. The affected region surrounding the missense variants (p.A349D and p.V436D) were aligned to other species and determined to be evolutionarily conserved. **(B)** Loss-of-function (LOF) OSMRβ variants reported in patients. To our knowledge, only the p.V436D variant is reported in gnomAD (https://gnomad.broadinstitute.org/), a large population database. The following in silico tools predicted the OSMRβ-LOF variants to be damaging, Combined Annotation Dependent Depletion (CADD) (https://cadd.gs.washington.edu/), Sorting Intolerant From Tolerant (SIFT) (https://sift.bii.a-star.edu.sg), and Polymorphism Phenotyping v2 (PolyPhen-2) (https://bio.tools/polyphen-2). SIFT and PolyPhen-2 cannot be applied to the p.Y660Sfs*16 variant, as it results in a premature stop codon and not an amino acid substitution.

### Unifying clinical features of the four patients with severe atopic disease

The four patients presented with a similar phenotype consistent with an underlying PAD (**Fig. 2**). Specifically, P2, P3 and P4 exhibited severe, widespread, treatment-resistant atopic dermatitis, which began in their first month of life combined with failure-to-thrive (**Fig. S1**) and atopic blood biomarkers of peripheral blood eosinophilia and markedly elevated serum IgE. P1 also had severe skin disease from early life, although it is notable that treatment with dupilumab resulted in a notable improvement in skin condition.

**Figure 2.**
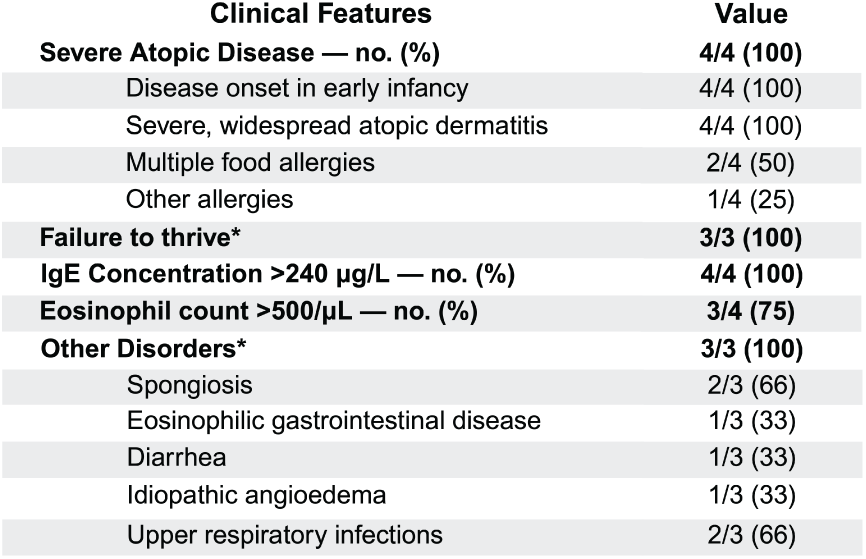
Major clinical features of the 4 patients. Tabulation and comparison of the clinical phenotype for 4 patients. *Please note for Patient 3, we had limited information about his family pedigree or childhood growth trajectory.

### p.A349D, p.V436D, and p.Y660Sfs*16 variants lead to a lack of OSMRβ cell surface expression

To assess the functional impact of the p.A349D, p.V436D, and p.Y660Sfs*16 OSMRβ variants, we modeled them in HEK293 cells. We selected HEK293 cells as our model system because these cells lack endogenous OSMRβ (Maier et al., 2015) (**Fig. 3A**). As controls, in parallel we tested WT OSMRβ and four OSMRβ variants reported in association with FPLCA (Arita et al., 2008; Lin et al., 2010; Tanaka et al., 2010; Wali et al., 2015). WT and variant OSMRβ proteins were tagged to a green-fluorescence protein (GFP) at the C-terminus, and over-expressed in the HEK293 cells. Using this system, we demonstrated significantly decreased cell surface expression of the p.A349D, p.V436D, and p.Y660Sfs*16 OSMRβ variant constructs compared to WT OSMRβ (*P* <.0001) (**Fig. 3A-D**). Interestingly, cell surface expression was intermediate in cells transfected with FPLCA variants (p.N462S, p.G513D, p.V631L, and p.I691T) (*P* <.0001). Notably, total OSMRβ protein expression was indistinguishable between all groups, as shown by intracellular flow cytometry staining (**Fig. 3E-F**). Additionally, using immunoblotting techniques, we established that when overexpressed following transfection, the p.Y660Sfs*16 variant leads to a truncated, stable OSMRβ protein (**Fig. 3G, Fig. S2A**), which is unable to translocate to the cell surface likely because the truncation removes the transmembrane domain of OSMRβ (**Fig. 1A**). Flow cytometry experiments validated that the p.Y660Sfs*16 variant results in a truncated OSMRβ protein that localizes to the intracellular compartment, as shown by high total OSMRβ expression, minimal extracellular OSMRβ expression, and no detectable expression of the protein’s GFP tag at its C-terminus. (**Fig. S2 B**). Together, these data indicate a mechanism for a LOF phenotype caused by the p.A349D, p.V436D, and p.Y660Sfs*16 OSMRβ variants, which all have distinctly lower cell surface expression patterns compared to both WT and FPLCA variants.

**Figure 3.**
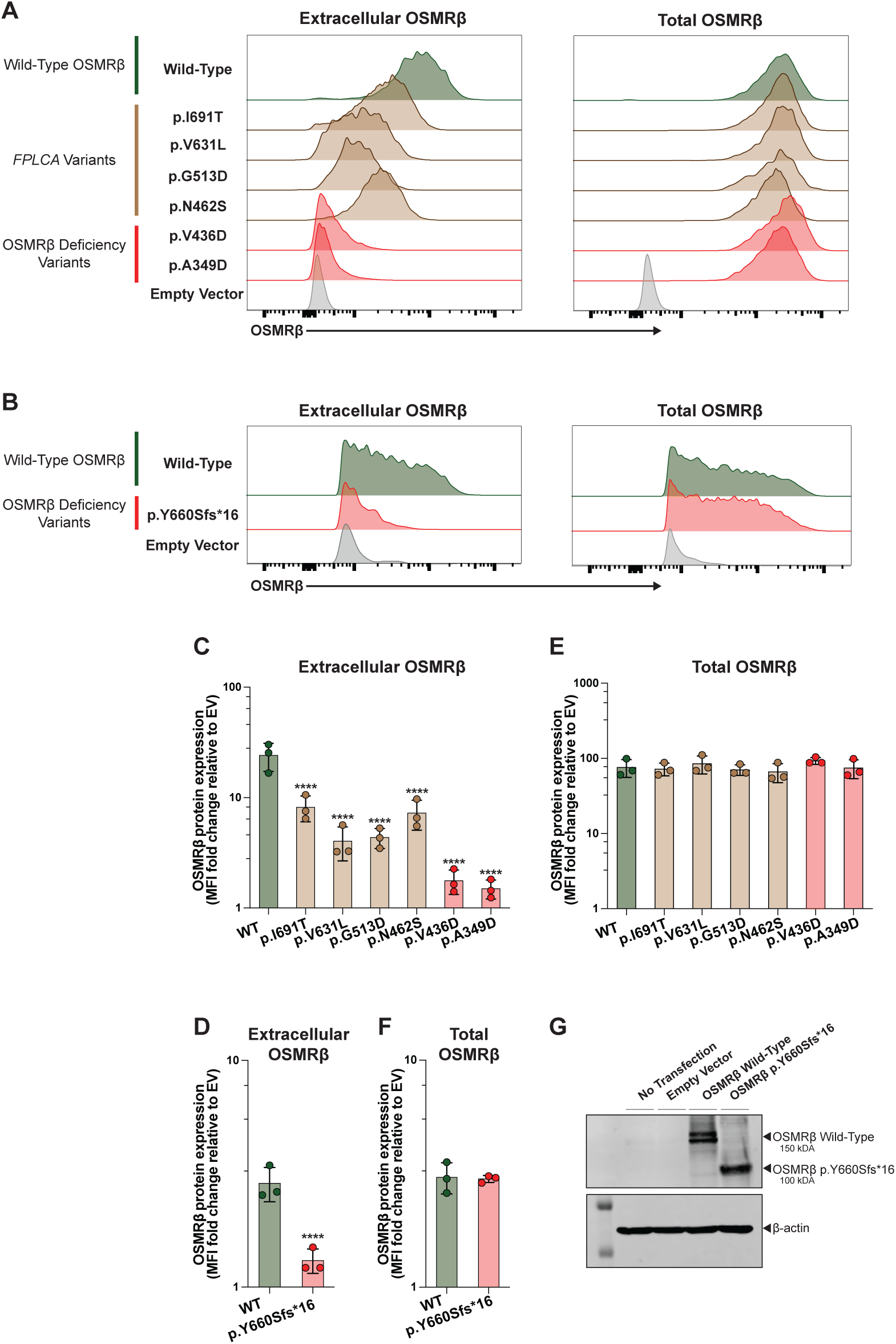
*OSMR*-LOF variants lead to a lack of OSMRβ cell surface expression in HEK293 cells. **(A)** Extracellular and total OSMRβ expression were quantified in HEK293 cells transfected with wild-type OSMRβ, OSMRβ deficiency variants, (p.A349D and p.V436D), or FPLCA variants (p.N462S, p.G513D, p.V631L, and p.I691T) using flow cytometry. OSMRβ expression was quantified in GFP^+^ cells. **(B)** Extracellular and total OSMRβ expression were quantified in HEK293 cells transfected with either wild-type OSMRβ or the OSMRβ deficiency variant p.Y660Sfs*16 variant using flow cytometry. OSMRβ expression was quantified in PE-OSMRβ^+^ cells instead of GFP^+^ cells as the variant results in a truncated protein, preventing translation of the C-terminal GFP. **(C)** Quantification of (A), extracellular OSMRβ expression, *n* = 3. Statistical comparisons based on one-way ANOVA and Dunnett’s multiple comparisons test. Stars denote *P* values: ****<.0001. **(D)** Quantification of (A), total OSMRβ expression, *n* = 3. Statistical comparisons based on unpaired T test. Stars denote *P* values: ****<.0001. **(E)** Quantification of (B), extracellular OSMRβ expression, *n* = 3. **(F)** Quantification of (B), total OSMRβ expression, *n* = 3. **(G)** Immunoblot visualizing OSMRβ expression in HEK293 cells transfected with wild-type OSMRβ or the p.Y660Sfs*16 variant; *n* = 3. The uncropped immunoblot can be found in Fig. S2 A.

### OSMRβ cell surface expression is reduced in primary fibroblasts

To validate the findings from the *in vitro* HEK293 cell experiments, we quantified OSMRβ expression in primary fibroblasts from P1, P2, and P3 alongside three healthy controls; primary samples from P4 were not available. Cell surface expression was significantly lower in the primary patient fibroblasts compared to healthy controls (*P* =.02) (**Fig. 4A-B**), whereas total OSMRβ expression was similar between patients and controls (**Fig. 4A**, **4C**). Notably, the cell surface OSMRβ expression in the healthy heterozygous parent of P1 (Family A-I-2) was comparable to the other two healthy controls, further reinforcing the autosomal recessive inheritance pattern of disease (**Fig. 4A**).

**Figure 4.**
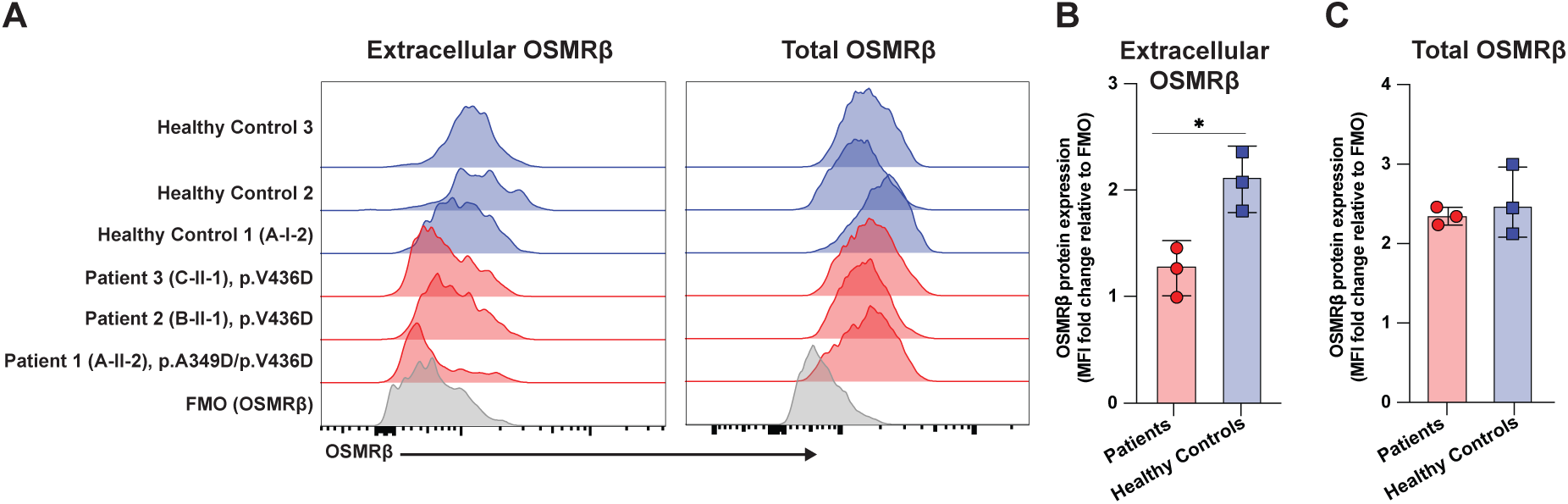
*OSMR* variants lead to decreased OSMRβ cell surface expression in primary fibroblasts. **(A)** Extracellular and total OSMRβ expression were quantified in primary fibroblasts from Patients 1, 2, and 3 (unable to obtain fibroblasts from Patient 4) and three healthy controls using flow cytometry. **(B)** Quantification of (A), extracellular OSMRβ expression, *n* = 3. Statistical comparisons based on unpaired t test. Stars denote *P* values: *<.05. FMO: fluorescence minus one.

### Biallelic LOF variants in *OSMR* impair signaling through the OSM/OSMRβ axis

Given that cell surface OSMRβ expression was low on primary cells, we hypothesized that this would likely impair signaling through the OSM/OSMRβ axis (**Fig. 5A**). We tested this hypothesis using primary fibroblasts from P1, P2, and P3. Activation of STAT5 after stimulation with OSM is exclusively mediated by the OSM type II receptor (OSMR/GP130), and was absent in patient cells. Similarly, patient cells displayed significantly decreased activation of STAT1 and STAT3 after OSM stimulation, with the residual activation likely reflecting signaling through the OSM type I receptor (LIFR/GP130). (**Fig. 5B-E** and **Fig. S3A**). Notably, heterozygous control fibroblasts from a parent (Family A-I-2) were indistinguishable from the healthy control fibroblasts, suggesting that impairment of the OSM/OSMRβ axis requires biallelic LOF. To establish that this signal defect was specific, we demonstrated that LIF signaling through the LIFR/GP130 complex was intact in all three patients (**Fig. 5F-H**, **Fig. S3B**) (Ichihara et al., 1997). Through these experiments in primary patient cells, we established that patients carrying biallelic LOF *OSMR* variants have impaired OSMRβ signaling while signaling through the LIFR/GP130 complex remained intact.

**Figure 5.**
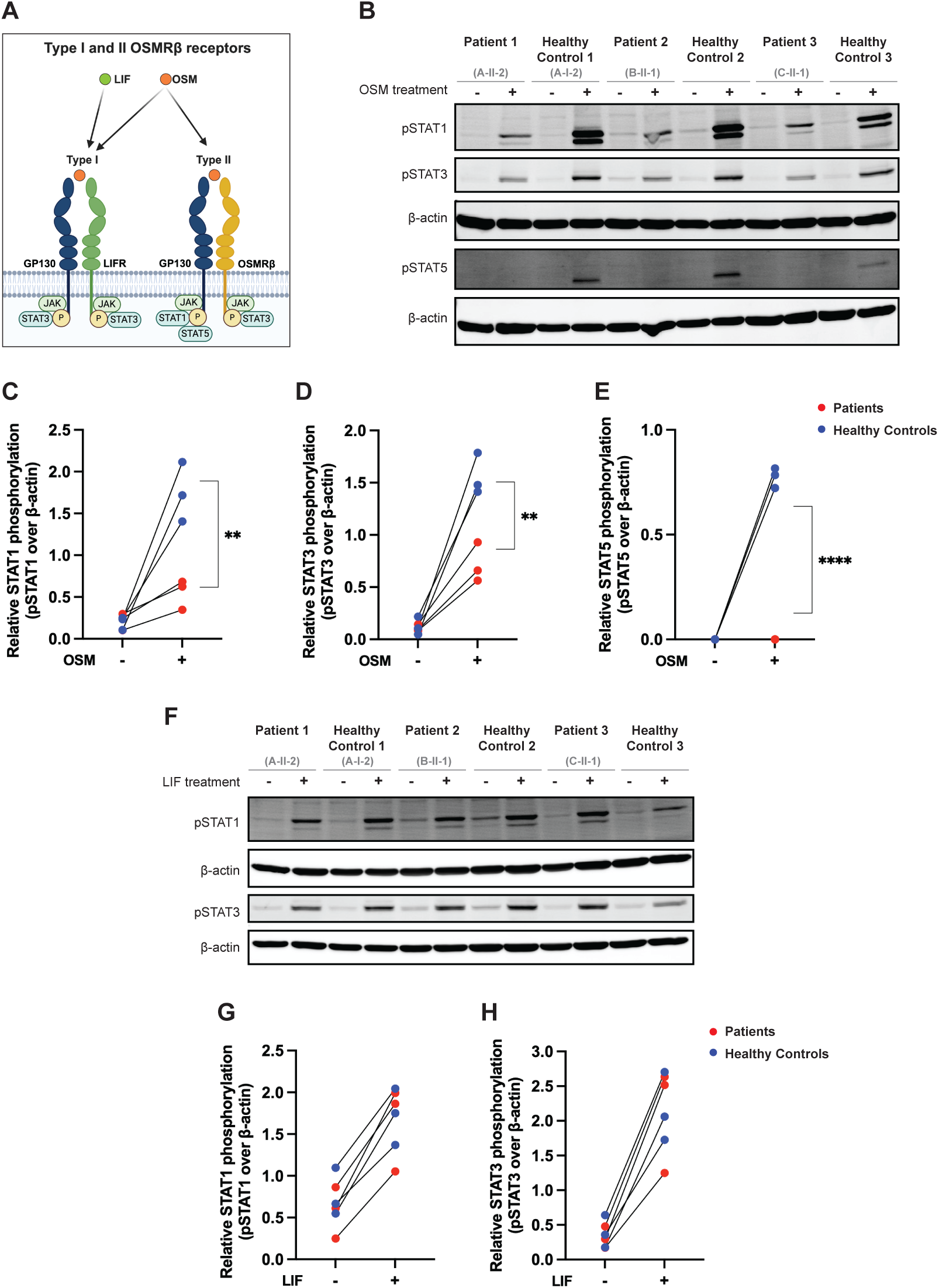
*S*elective impairment of STAT signaling through the OSM/OSMRβ axis in primary fibroblasts. **(A)** Schematic illustrating the OSM/OSMRβ axis. **(B)** Immunoblot in primary fibroblasts from Patients 1, 2, and 3 (unable to obtain fibroblasts from Patient 4) and three healthy controls for pSTAT1, pSTAT3, and pSTAT5 before and after treatment with OSM (100ng/ml for 15 minutes); *n* = 3. Full-length immunoblot for this can be found in Fig. S3 A. **(C-E)** Quantification of the immunoblot in (B) for **(C)** pSTAT1, **(D)** pSTAT3, and **(E)** pSTAT5. Patients are in red and healthy controls are in blue. Statistical comparisons based on unpaired t test. Stars denote *P* values: **, *P* <.01; ****, *P* <.0001. **(F)** Immunoblot in primary fibroblasts from Patients 1, 2, and 3 and three healthy controls for pSTAT1 and pSTAT3 before and after treatment with LIF (100ng/ml for 15 minutes); *n* = 3. Full-length immunoblot for this can be found in Fig. S3 B. **(G-H)** Quantification of the immunoblot in (F) for **(G)** pSTAT1 and **(H)** pSTAT3. Patients are in red and healthy controls are in blue.

### Expression of WT OSMRβ restores OSM signaling in patient fibroblasts

To establish a causal relationship between the genotype and cellular phenotype (Casanova et al., 2014), we used a lentiviral approach to re-express WT OSMRβ in primary dermal fibroblasts of P2. Lentiviral transduction rescued cell surface expression of OSMRβ (**Fig. 6A-C**). Lentiviral transduction also rescued signaling through the OSM axis, as measured by pSTAT1 and pSTAT3 (**Fig. 6D**, **Fig. 6E**, **Fig. 6F**, **Fig. S3C**, and **Fig. S4**). Finally, analysis of transcriptomic signatures confirmed that most of the differentially regulated genes in healthy controls were absent in P2 fibroblasts but were rescued in WT *OSMR*-transduced fibroblasts (**Fig. 6G-H**). Healthy control fibroblasts had 523 significantly upregulated genes upon stimulation with OSM and 211 downregulated ones. We observed no differential gene expression in P2 fibroblasts (EV) after treatment. We observed 91 upregulated genes in P2 fibroblasts (rescued) and no downregulated genes, among which 85 overlapped with upregulated genes seen in healthy control fibroblasts (**Fig. 6H**).

**Figure 6.**
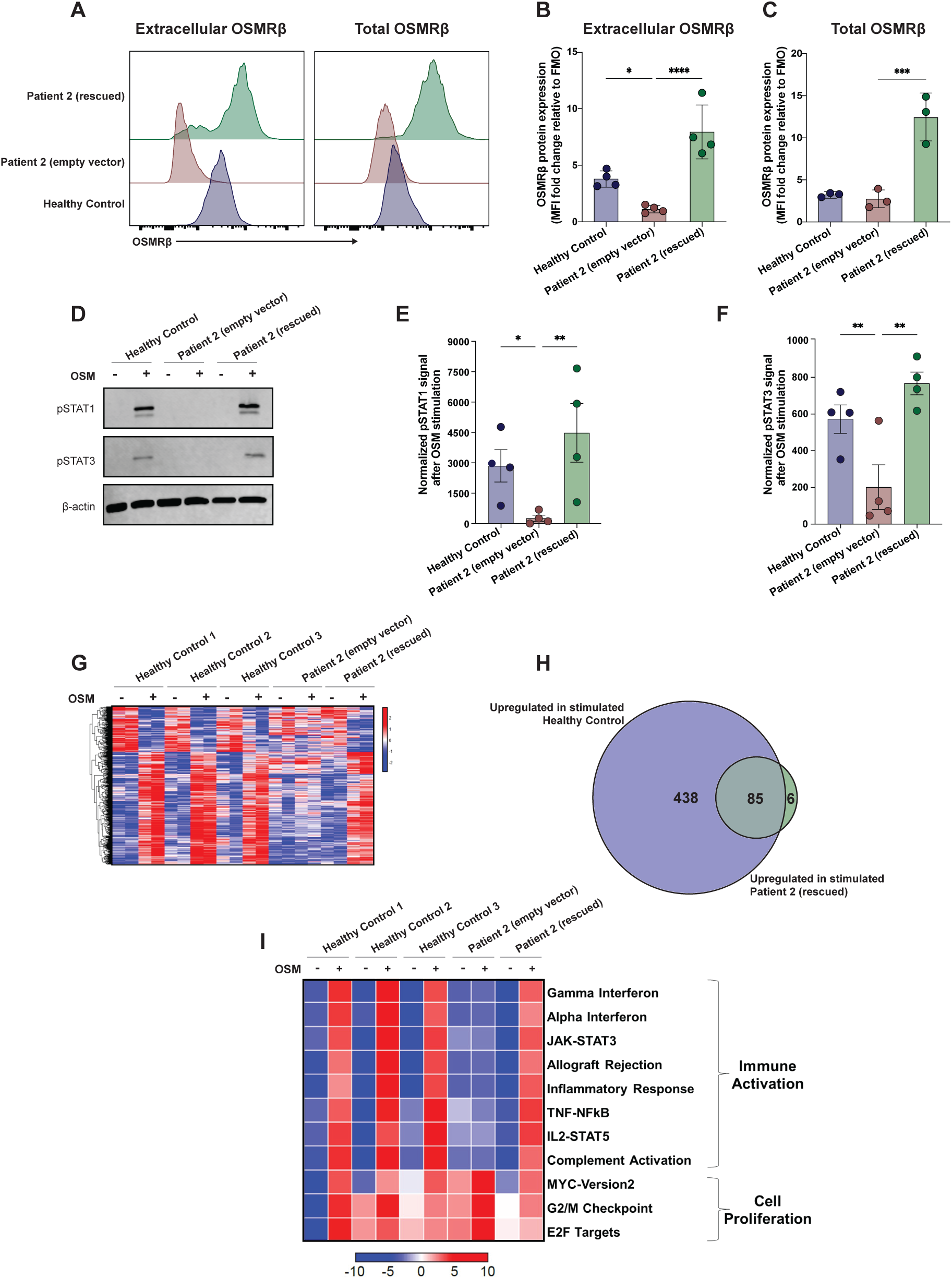
WT-*OSMR* restores the OSM/OSMRβ axis and STAT signaling in primary fibroblasts. **(A)** Extracellular and total OSMRβ expression were quantified in primary fibroblasts from Patient 2 that were transduced with WT-*OSMR* (rescued) or an empty vector and compared to primary fibroblasts from a healthy control using flow cytometry. **(B)** Quantification of (A), extracellular OSMRβ expression, *n* = 3. **(C)** Quantification of (A), total OSMRβ expression, *n* = 3. **(D)** Immunoblot in primary fibroblasts from Patient 2 that were transduced with WT-*OSMR* or an empty vector and compared to primary fibroblasts from a HC for pSTAT1 and pSTAT3 before and after treatment with OSM (100ng/ml for 15 minutes); *n* = 4. Full-length immunoblot can be found in Fig. S3 C. **(E-F)** Quantification of the immunoblot in (D) for **(E)** pSTAT1 and **(F)** pSTAT3. **(G)** Heatmap signatures of differentially expressed genes comparing P2 fibroblasts (either transduced with WT-*OSMR* or an empty vector) against three HCs before and after stimulation with OSM (100ng/ml for 15 minutes). **(H)** Overlap in significantly upregulated genes upon OSM stimulation in HC cells (blue) and P2 rescued cells (green) as shown through a Venn diagram. **(I)** Gene set enrichment analysis (GSEA) of significantly enriched immune pathways from the MSigDB Hallmark in P2 (either transduced with WT-*OSMR* or an empty vector) against three HCs before and after stimulation with OSM (100ng/ml for 15 minutes). Heatmap is normalized across the rows and shown as relative expression of sample level enrichment scores. Statistical comparisons based on one-way ANOVA and Dunnett’s multiple comparisons test. Stars denote *P* values: *<.05, **<.01, ***<.001, and ****<.0001.

Differential pathway activation using gene set enrichment analysis (GSEA) showed significant enrichment in pathways for immune activation and cell proliferation in healthy controls after stimulation with OSM (**Fig. 6I**). Consistent with established links between STAT1 activation and interferon signaling (Dupuis et al., 2001; Dupuis et al., 2003), we found that OSM stimulation led to strong activation of interferon activation pathways (e.g., healthy control (HC): Gamma_IFN NES 2.71, adj *P*-value <.001), as well as STAT3, STAT5 and MYC pathway activation (**Fig. 6I, Table S2).** These pathway activations were either reduced or absent in patient fibroblasts (**Fig. 6I, Table S2)**. Reinforcing causality, lentiviral rescue of P2 fibroblasts with WT-*OSMR* restored signaling in all immune-activated pathways. Collectively, these findings establish the role of OSMRβ in OSM-mediated immune activation of fibroblasts.

## DISCUSSION

In this study, we present a combination of clinical, genetic, molecular, and transcriptional evidence defining a new human PAD caused by biallelic germline LOF *OSMR* variants in four patients with early-onset severe atopic disease. These variants all led to a lack of cell surface expression of OSMRβ protein and decreased OSM-mediated STAT1, STAT3, and STAT5 activation and phosphorylation. Given that we are only describing 4 patients, it should be emphasized that the full phenotype of biallelic *OSMR* LOF will only be resolved through the identification of additional affected individuals.

Population genetics provide another layer of evidence supporting the involvement of genetic variation of *OSMR* in human immunity. Several independent genome-wide association studies (GWAS) have found that polymorphisms in *OSMR* associate with multiple chronic inflammatory conditions, including Crohn’s disease (de Lange et al., 2017; Ellinghaus et al., 2016; Liu et al., 2015), ulcerative colitis (de Lange et al., 2017; Ellinghaus et al., 2016; Liu et al., 2015), psoriasis (Ellinghaus et al., 2016), ankylosing spondylitis (Ellinghaus et al., 2016), and sclerosing cholangitis (Ellinghaus et al., 2016). Interestingly, polymorphisms in *OSMR* also associate with the measurement of von Willebrand factor (Temprano-Sagrera et al., 2022), a blood clotting protein. Deficiency of this protein leads to von Willebrand disease, which was diagnosed in P1. Recently, microbial genome-wide interaction studies found that interactions between single-nucleotide polymorphisms in the *OSMR* gene and early-life exposures, such as breastfeeding, are associated with gut microbiota implicated in asthma and atopy (Stickley et al., 2025).

An important distinction we establish in this study is the difference between biallelic autosomal recessive OSMRβ deficiency and FPLCA. Whilst there is overlap between the conditions, there are key differences. The most notable are that FPLCA is typically an autosomal dominant disorder with disease limited to the skin and onset in adolescence or adulthood, while the patients we describe all had the onset of severe atopic dermatitis from infancy accompanied by other manifestations of their allergic diathesis. Moreover, the amyloid deposits in the superficial dermis, which are a defining feature of FPLCA, were lacking in the skin biopsies of patients with biallelic OSMRβ deficiency (Tanaka et al., 2010). Functional data emphasize the differences between FPLCA and biallelic OSMRβ deficiency. We show that FPLCA variants lead to intermediate levels of cell surface expression of OSMRβ which contrasts with the absence of cell surface expression we find with the autosomal recessive *OSMR*-LOF variants. Our result is consistent with a previous study of OSMRβ expression by microscopy that found no differences in the localization of FPLCA-associated OSMRβ variants when compared to OSMRβ^WT^ (Liu et al., 2021). Ultimately, the distinct features and potential overlap between autosomal dominant FPLCA and autosomal recessive OSMRβ deficiency will be fully defined through the diagnosis of additional affected individuals. None of our identified variants have been associated with FPLCA, despite multiple clinical reports of this condition.

While mouse models can be powerful tools, we must be cautious in extrapolating the biology of the human OSM/OSMRβ axis from mouse studies. Mouse Osm shares only 48% amino acid identity with human OSM (Yoshimura et al., 1996), and mouse Osm only activates the OSMRβ/GP130 receptor complex, whereas human OSM can activate both the LIFR/GP130 and the OSMRβ/GP130 receptor complexes (Ichihara et al., 1997). As such, comparing the patients we describe with biallelic OSMRβ deficiency with *Osmr^-/-^* mouse comes with several caveats. Nevertheless, it is notable that *Osmr*^−/−^ mice do have a skin phenotype with an increase in tail epidermal thickness (Liu et al., 2021). However, other phenotypes of *Osmr^-/-^* mice, including thrombocytopenia and anemia (Tanaka et al., 2003), impaired acute inflammation (Lorchner et al., 2015), and abnormalities in bone development (Guihard et al., 2015; Walker et al., 2010), were not noted in patients with autosomal recessive OSMRβ deficiency. These differences likely result in non-redundant, non-overlapping signalling in these linked pathways. Indeed, Stüve-Wiedemann syndrome type 1 in humans — caused by autosomal recessive *LIFR* LOF — shares features of skeletal abnormalities but not others (Dagoneau et al., 2004). In contrast, Stüve-Wiedemann syndrome type 2 — caused by recessive IL6ST LOF encoding GP130 — also has features of bone abnormalities, in addition to thrombocytopenia, atopic dermatitis, and impaired immunity (Schwerd et al., 2017).

The discovery of IEIs has a rich tradition of informing our understanding of fundamental human biology, and some valuable insights can be gained from the discovery of these patients with autosomal recessive OSMRβ deficiency. We observed that OSM-mediated OSMRβ activation leads to strong STAT3-dependent signaling in dermal fibroblasts, suggesting that the OSM/OSMRβ axis might play a central role in stromal cell STAT3 activation. Indeed, OSM is one of the strongest activators of STAT3 *in vitro*, and thus has been used to assess the functional impact of variants affecting this pathway (Chandrasekaran et al., 2016). This knowledge underscores the potential mechanism behind “cold abscesses”, lack of skin inflammation, and cutaneous infections seen in *DN*-STAT3 HIES patients (Tsilifis, Freeman, and Gennery, 2021). We further propose that biallelic *OSMR* disruption could be linked to enhanced IL-4/STAT6 activation, possibly leading to barrier disruption (Strid, McLean, and Irvine, 2016) (Sharma et al., 2024), since P1’s severe atopic dermatitis dramatically improved after dupilumab (anti-IL-4Rα antibody) treatment. Previous studies have demonstrated that small deletions in GP130 result in a decrease in cell surface expression while causing ligand-independent intracellular signaling (Rinis et al., 2014), which can be targeted by JAK inhibitors (Poussin et al., 2013).

We want to explicitly acknowledge that one of the *OSMR* variants that we report (*OSMR* c.1307T>A, p.V436D) is relatively common in publicly available databases with 5 homozygotes of European ancestry reported in gnomAD v4.1.0 (non-UKB). While the functional data we report for this variant are compelling, this suggests that *OSMR*-LOF might be a relatively frequent cause of severe allergic disease. There is precedent for this in other monogenic allergic disorders. Specifically, relatively common filaggrin (*FLG*) variants are associated with atopic dermatitis and secondary allergic diseases (Sandilands et al., 2007), and more recently, *JAK1* variants with mild-to-moderate gain-of-function activity were linked to common presentations of inflammatory, allergic, and/or autoimmune diseases (Horesh et al., 2024). It is notable that an increased prevalence of recurrent viral and bacterial infections has been reported in a subset of children with severe atopic dermatitis (Ong and Leung, 2016). While certainly not all of these patients will have underlying *OSMR* LOF variants, the pathways and mechanisms we identify could be generalizable to this endotype of severe allergic disease and recurrent viral infections, perhaps opening novel therapeutic avenues.

In conclusion, this first description of patients with biallelic LOF variants in *OSMR* causing a new PAD expands our understanding of the role of OSMRβ in human health and disease. Given that now at least 50 genes have been recognized to cause PADs (Vaseghi-Shanjani et al., 2025), we encourage clinicians to employ inclusive next-generation sequencing approaches to pursue a diagnosis of PADs in their patients with unusual and severe allergic disease since timely diagnosis and molecularly-targeted treatments can be transformative for affected individuals.

## MATERIALS AND METHODS

### Ethical Considerations

All study participants and/or their parents/guardians provided written informed consent. Research study protocols were approved by The University of British Columbia Clinical Research Ethics Board (H15-0064), or the NIH Institutional Review Board (NCT00852943, NCT01164241).

### Identification of *OSMR* variants using whole-exome sequencing

Trio whole-exome sequencing (WES) was performed on genomic DNA from all patients while siblings were assessed for segregation of variants. The identified *OSMR* variants were predicted to be damaging and segregated with disease and thus were selected for further analysis (Rentzsch et al., 2021) (Sim et al., 2012) (Adzhubei et al., 2010).

### Generation and expression of *OSMR* variant plasmids

*OSMR* variants described in this study were generated through site-directed mutagenesis using primer pairs noted in Table S1 for transfection purposes. Expression of wild-type (WT) OSMRβ or OSMRβ variants were induced transiently in HEK293 cells using lipofectamine, or stably in fibroblasts using a lentiviral approach. See supplemental methods at the end of the PDF for details.

### Isolation and culture of primary dermal fibroblasts

Primary fibroblasts were isolated from a non-lesional skin punch biopsy (P1 and P2) or a surgical skin biopsy (P3) and cultured at 37°C until a confluent monolayer of fibroblasts had formed. Fibroblasts were Sanger sequenced to confirm the genotype of the cells. See supplemental methods at the end of the PDF for details.

### Flow cytometry

(a) Extracellular flow cytometry: Presence of OSMRβ on the cell surface of viable, transfected HEK293 cells and primary fibroblasts was determined using an anti-OSMRβ antibody. (b) Intracellular flow cytometry: Total OSMRβ (extracellular and intracellular) in transfected HEK293 cells and primary fibroblasts was determined using an anti-OSMRβ antibody on fixed and permeabilized cells. (c) Phospho-flow cytometry: STAT1, STAT3, and STAT5 phosphorylation was quantified using phospho-flow cytometry on fixed and permeabilized fibroblasts from P2 and a healthy control. See supplemental methods at the end of the PDF for details.

### Immunoblotting

Immunoblotting was conducted as previously described (Samra et al., 2024; Sharma et al., 2023) to assess the phosphorylation status of STAT1, STAT3, and STAT5 in primary fibroblasts stimulated with OSM, IL-31, or LIF and to quantify OSMRβ expression in HEK293 cells. See supplemental methods at the end of the PDF for details.

### RNA Sequencing

To profile the global transcriptome, primary fibroblasts from three healthy controls and P2 were transduced with either empty vector (EV) or WT *OSMR*. Cell were stimulated with 100 ng/ml OSM for 4 hours or left unstimulated. RNA was extracted in duplicate and sequenced as previously described (Lu et al., 2021; Sharma et al., 2022; Sharma et al., 2023). See supplemental methods at the end of the PDF for details.

### Data Availability

Data is deposited at Gene Expression Omnibus (GEO) with the following accession number: GSE303962.

### Online Supplemental Material

Fig S1 contains growth charts for two out of the four patients. Fig S2 contains full-length immunoblot showing truncated p.Y660Sfs*16 OSMRβ variant and contour plots showing that the *OSMR* variants lead to a lack of OSMRβ cell surface expression in HEK293 cells. Fig S3 contains full-length immunoblots showing selective impairment of STAT signaling in primary fibroblasts, which is restored with WT-*OSMR* transduction. Fig S4 contains additional data showing that WT-*OSMR* transduction restores STAT signaling in primary fibroblasts. Table S1 lists primers used for site-directed mutagenesis. Table S2 list pathways that are significantly activated in patient and healthy control fibroblasts after stimulation with OSM.

## ACKNOWLEDGMENTS

We thank all the patients and their families for taking part in this study. This work was supported by grants from the Canadian Institutes of Health Research (PJT-178054; S.E. Turvey), Genome British Columbia (SIP007; S.E. Turvey), the Jeffery Modell Foundation (S.E. Turvey, J.J. Lyons), NIAID (4R00AI138586, J.J. Lyons), and NIH (DK062370, M. Boehnke). This research was supported in part by the Intramural Research Program of the National Institutes of Health (NIH). The contributions of the NIH author(s) were made as part of their official duties as NIH federal employees, are in compliance with agency policy requirements, and are considered Works of the United States Government. However, the findings and conclusions presented in this paper are those of the author(s) and do not necessarily reflect the views of the NIH or the U.S. Department of Health and Human Services. S.E. Turvey holds a Tier 1 Canada Research Chair in Pediatric Precision Health and the Aubrey J. Tingle Professor of Pediatric Immunology. S. Samra is supported by the University of British Columbia Four Year Doctoral Fellowship. We also thank Daniel MacArthur of the Broad Institute who through the creation of ExAC and gnomAD helped facilitate this collaboration.

## LIST OF ABBREVIATIONS

CADD: Combined Annotation Dependent Depletion
EV: Empty vector
GFP: Green fluorescent protein
GSEA: Gene set enrichment analysis
FPLCA: Familial primary localized cutaneous amyloidosis
IEI: Inborn errors of immunity
IL: Interleukin
LIF: Leukemia inhibitory factor
LOF: Loss-of-function
NES: Normalized enrichment scores
OSM: Oncostatin M
OSMRβ: Oncostatin M receptor beta
SIFT: Sorting Intolerant From Tolerant
PAD: Primary atopic disorder
PolyPhen: Polymorphism Phenotyping
WES: Whole-exome sequencing
WT: Wild-type

## SUPPLMENTARY MATERIALS AND METHODS

### Generation of *OSMR* variant plasmids

Plasmids used for transfection studies contained full-length *OSMR* cDNA in a pCMV6-AC-GFP vector with a C-terminal GFP tag (Cat#: RG216943, OriGene Technologies). To generate *OSMR* variants, c.1046C>A, c.1307T>A, c.1979_1980delAC, and other literature reported FPLCA variants, a Q5 site-directed mutagenesis kit (Cat#: E0554S, New England Biolabs) was used according to the manufacturer’s recommendations, with primer pairs noted in Table S1.

To generate lentivirus vectors, WT OSMRβ from the above plasmids were cloned into a Lenti vector with a C-terminal GFP tag (Cat#: PS100071, OriGene Technologies) using EcoRI-HF (Cat#: R3101, New England Biolabs) and NotI-HF (Cat#: R3189, New England Biolabs). The plasmids were packaged using third-generation packaging plasmids and transfected into HEK293T cells. This was also done for an empty Lenti vector which was used as a control. Culture media was collected, centrifuged, filtered, concentrated, and stored at −80°C before use.

All *OSMR* variant plasmids were confirmed by Sanger sequencing and purified from 10-beta competent *E. coli* using a QIAprep Spin Miniprep Kit (Cat#: 27104, Qiagen).

### Isolation and culture of primary dermal fibroblasts

Primary fibroblasts were isolated from a non-lesional skin punch biopsy (P1 and P2) or a surgical skin biopsy (P3). The skin was excised from the underlying connective tissue and placed in separate wells of a 6-well plate in DMEM (GE Healthcare) supplemented with 10% heat-inactivated FBS (Gibco, Life Technologies), 2mM L-glutamine (HyClone, Thermo Fisher Scientific), 1mM sodium pyruvate (Gibco, Life Technologies), and 1x Antibiotic-Antimycotic (Gibco, Thermo Fisher) for 3 weeks at 37°C, or until a confluent monolayer of fibroblasts had formed. Primary fibroblasts were then lifted and frozen for future assays.

### Transient and stable expression of *OSMR* variants

Transient expression of *OSMR* variants in HEK293 cells was accomplished using a Lipofectamine 3000 kit (Thermo Fisher Scientific) according to the manufacturer’s recommendation. Briefly, HEK293 cells were seeded at 8.0 × 10^5^ cells/well in a 6-well plate in 1.5 mL of DMEM supplemented with 10% FBS and incubated for 24 hours at 37°C. Cells were transfected with 2.5 μg of plasmid DNA using P3000 and Lipofectamine 3000 reagents and harvested after 24h.

Stable expression of OSMRβ in primary dermal fibroblasts was accomplished using the lentivirus approach as previously described (Fung et al., 2021; Kutner, Zhang, and Reiser, 2009). Briefly, primary dermal fibroblasts were infected with lentiviral particles in the presence of 5 μg/ml polybrene (Sigma-Aldrich), cultured, and expanded in DMEM supplemented with 10% FBS (Gibco, Life Technologies). Expanded cells were sorted on GFP expression using a BD FACS Aria (BD Biosciences) cell sorter.

### Cell surface and intracellular flow cytometry

OSMRβ expression and phospho-STAT detection were quantified using flow cytometry in transfected and primary cells. For cell surface OSMRβ expression, transfected HEK293 cells or primary fibroblasts were lifted and stained with *OSMR* PE (Cat#:12-1303-42, ThermoFisher) for 20 mins. Expression was measured in GFP^+^ HEK293 cells or primary fibroblasts on an LSRII flow cytometer (BD Biosciences) and analyzed using FlowJo software (BD Biosciences). For total OSMRβ and phospho-STAT detection, cells were fixed using BD Cytofix (Cat#: 554655, BD Biosciences) for 20 min at 4°C and permeabilized using Perm III for 30 min on ice (Cat#: 558050, BD Biosciences). The cells were then stained with *OSMR* PE, or with pSTAT1 BV421 (Cat#: 562985, BD Biosciences), pSTAT3 PECF594 (Cat#: 562673, BD Biosciences), and pSTAT5 PeCy7 (Cat#: 560117, BD Biosciences). OSMRβ expression was measured in GFP^+^ HEK293 cells that were transfected with the p.A349D and p.V436D variants, PE^+^ HEK293 cells that were transfected with p.Y660Sfs*16 variant (because this variant results in a truncated protein and thus the C-terminal GFP would not be translated), and GFP^+^ primary fibroblasts. Cells were analyzed on an LSRII flow cytometer (BD Biosciences), and flow cytometry data were analyzed using FlowJo software (BD Biosciences).

### Immunoblotting

The phosphorylation status of STAT1, STAT3, and STAT5 was determined in primary fibroblasts cultured in DMEM. OSMRβ expression was measured in HEK293 cells cultured in DMEM. Cells were harvested in chilled RIPA buffer (Cat# 89901; Thermo Fisher Scientific) supplemented with HALT protease and phosphatase inhibitor cocktail (Cat# 87786; Thermo Fisher Scientific) and then lysed for 15 minutes on ice. Cell lysates were separated by 12% SDS-polyacrylamide gel electrophoresis and transferred onto PVDF membranes (Cat# IPFL00010; Immobilon-FL; MilliporeSigma). Membranes were blocked with 5% BSA in TRIS-buffered saline with Tween-20 for an hour and then incubated with primary antibodies in blocking buffer overnight at 4°C. The next day, membranes were washed and incubated with secondary antibodies for 1 hour at room temperature and then imaged using a LI-COR Odyssey infrared scanner (LI-COR Biosciences).

The primary antibodies used were the following: pSTAT1 (Cat#: 9167, Cell Signaling Technologies), pSTAT3 (Cat#: 9138, Cell Signaling Technologies), pSTAT5 (Cat#: 9356, Cell Signaling Technologies), OSMRβ (ab282577, Abcam), and β-actin (Cat#: 3700, Cell Signaling Technologies). The secondary antibodies used were the following: goat anti-rabbit IgG DyLight 800 conjugated (Cat#: 611-145-002-0.5, Rockland Immunochemicals) and goat anti-mouse IgG IRDye 680RD (Cat#: 926-6870, LI-COR).

### RNA sequencing

RNA was extracted, sequenced, and then pre-processed as previously described (Lu et al., 2021; Sharma et al., 2022; Sharma et al., 2023). Expression data were then normalized to reads between samples using the edgeR package in R (R Foundation). Normalized counts were filtered to remove low counts using the filterByExpr function in edgeR (Chen, Lun, and Smyth, 2016). Differential expression between unstimulated and stimulated samples for healthy control fibroblasts, P2-EV (empty vector) fibroblasts, and P2-WT (wild type)-*OSMR* was accomplished using Limma (Ritchie et al., 2015). Differentially expressed genes were defined as those with a fold change greater than 1.25 and an Benjamini-Hochberg adjusted *P*-value less than 0.05.

Pathway analysis was done by first performing GSEA with 1000 permutations using the Molecular Signatures Database Hallmark module. The signal-to-noise ratio was used for gene ranking and the obtained normalized enrichment scores (NES), and P-values were further adjusted using the Benjamini-Hochberg method. Pathways with an adjusted *P*-value <.05 were considered significant. Leading-edge genes from significant pathways between healthy control unstimulated fibroblasts and stimulated fibroblasts were identified. Expression levels of these genes were then determined in each of the three groups (healthy control fibroblasts, P2-EV and P2-WT-*OSMR*). Sample level enrichment analyses scores were computed as previously described (Kulpa et al., 2019). Briefly, z-scores were computed for gene sets of interest for each sample. The mean expression levels of significant genes were compared to the expression of 1000 random gene sets of the same size. The difference between observed and expected mean expression was then calculated and represented on heatmaps.

---

**Supplementary Figure 1.**
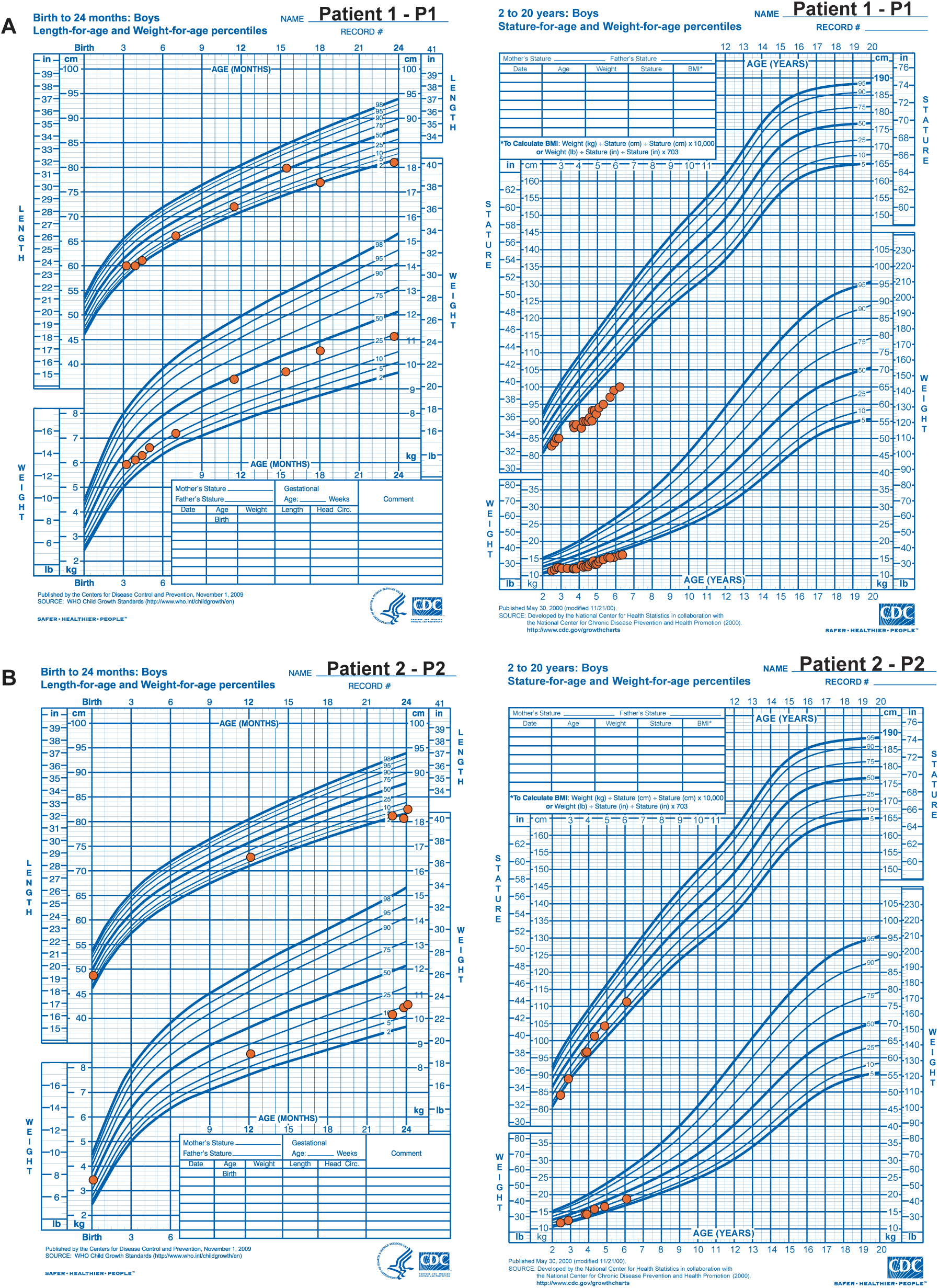
Growth charts for **(A)** Patient 1 and **(B)** Patient 2.

**Supplementary Figure 2.**
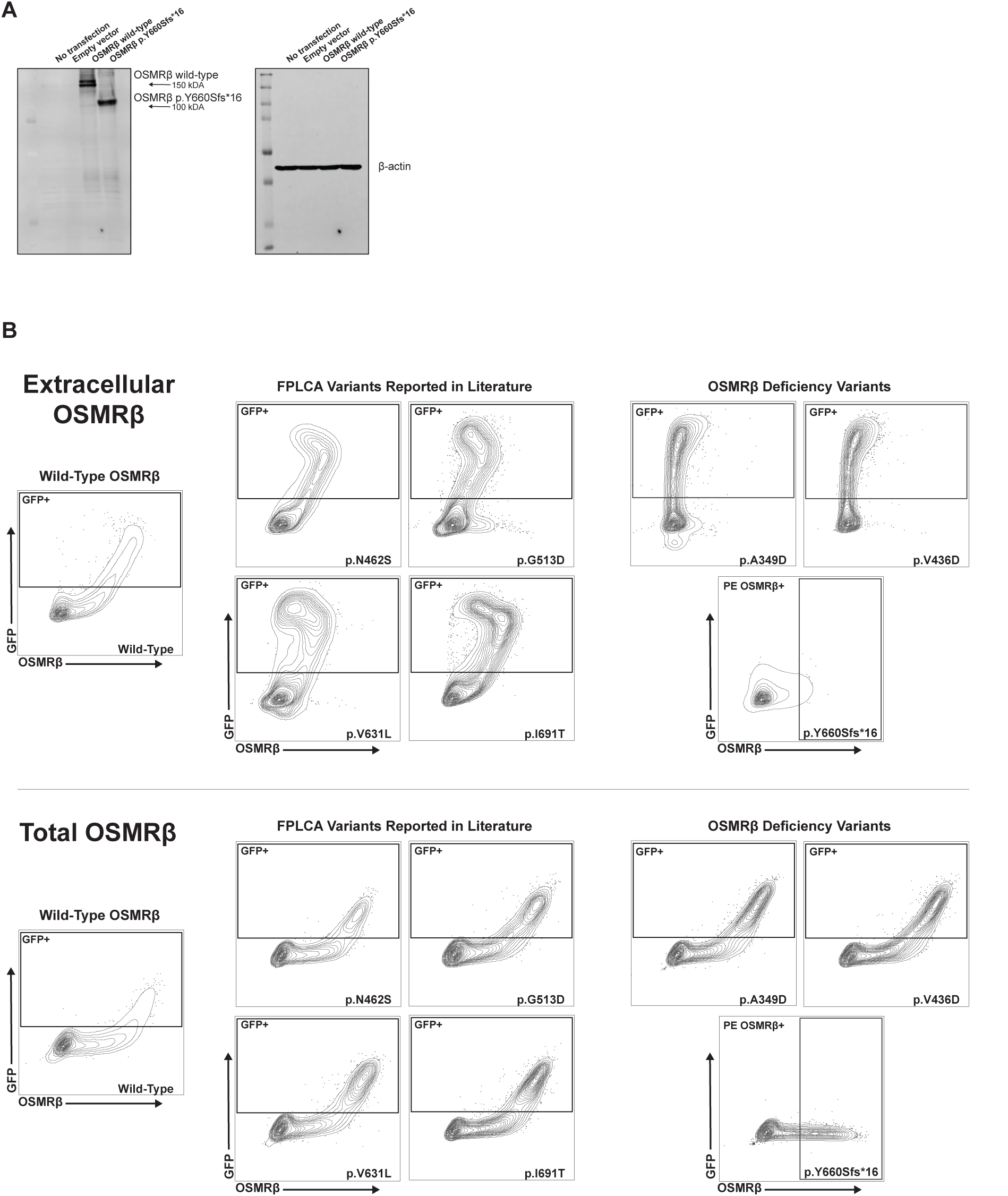
Full-length immunoblot showing truncated p.Y660Sfs*16 OSMRβ variant and contour plots showing that *OSMR*-LOF variants lead to a lack of OSMRβ cell surface expression in HEK293 cells. **(A)** Full-length immunoblots of the cropped immunoblots shown in Fig. 3 G. Immunoblots in HEK293 cells transfected with OSMRβ (wild-type or the p.Y660Sfs*16 variant) or an empty vector control; *n* = 3. **(B)** Extracellular and total OSMRβ expression were quantified in HEK293 cells transfected with wild-type OSMRβ, OSMRβ LOF variants, (p.A349D, p.V436D, and p.Y660Sfs*16), or FPLCA variants (p.N462S, p.G513D, p.V631L, and p.I691T) using flow cytometry.

**Supplementary Figure 3.**
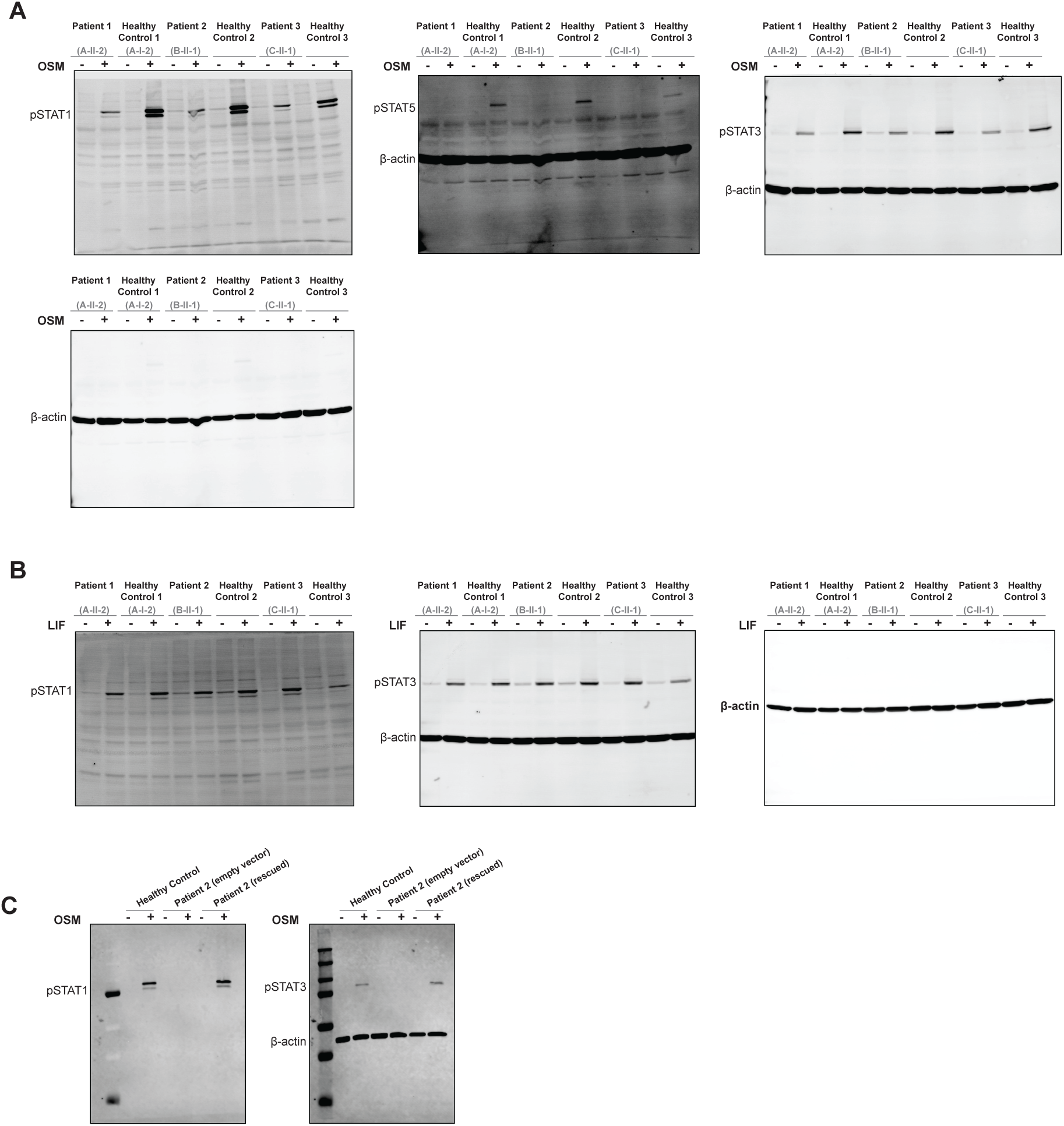
Full-length immunoblots showing selective impairment of STAT signaling through the OSM/OSMRβ axis in primary fibroblasts, which is restored with WT-*OSMR* transduction. **(A)** Full-length immunoblots of the cropped immunoblots shown in Fig. 5 B. Immunoblots in primary fibroblasts from Patients 1, 2, and 3 alongside three healthy controls for pSTAT1, pSTAT3, and pSTAT5 before and after treatment with OSM (100ng/ml for 15 minutes); *n* = 3. **(B)** Full-length immunoblots of the cropped immunoblots shown in Fig. 5 F. Immunoblots in primary fibroblasts from Patients 1, 2, and 3 alongside three healthy controls for pSTAT1 and pSTAT3 before and after treatment with LIF (100ng/ml for 15 minutes); *n* = 3. **(C)** Full-length immunoblots of the cropped immunoblots shown in Fig. 6 D. Immunoblot in primary fibroblasts from Patient 2 that were either transduced with WT-*OSMR* or an empty vector and compared to primary fibroblasts from a healthy control for pSTAT1 and pSTAT3 before and after treatment with OSM (100ng/ml for 15 minutes); *n* = 4.

**Supplementary Figure 4.**
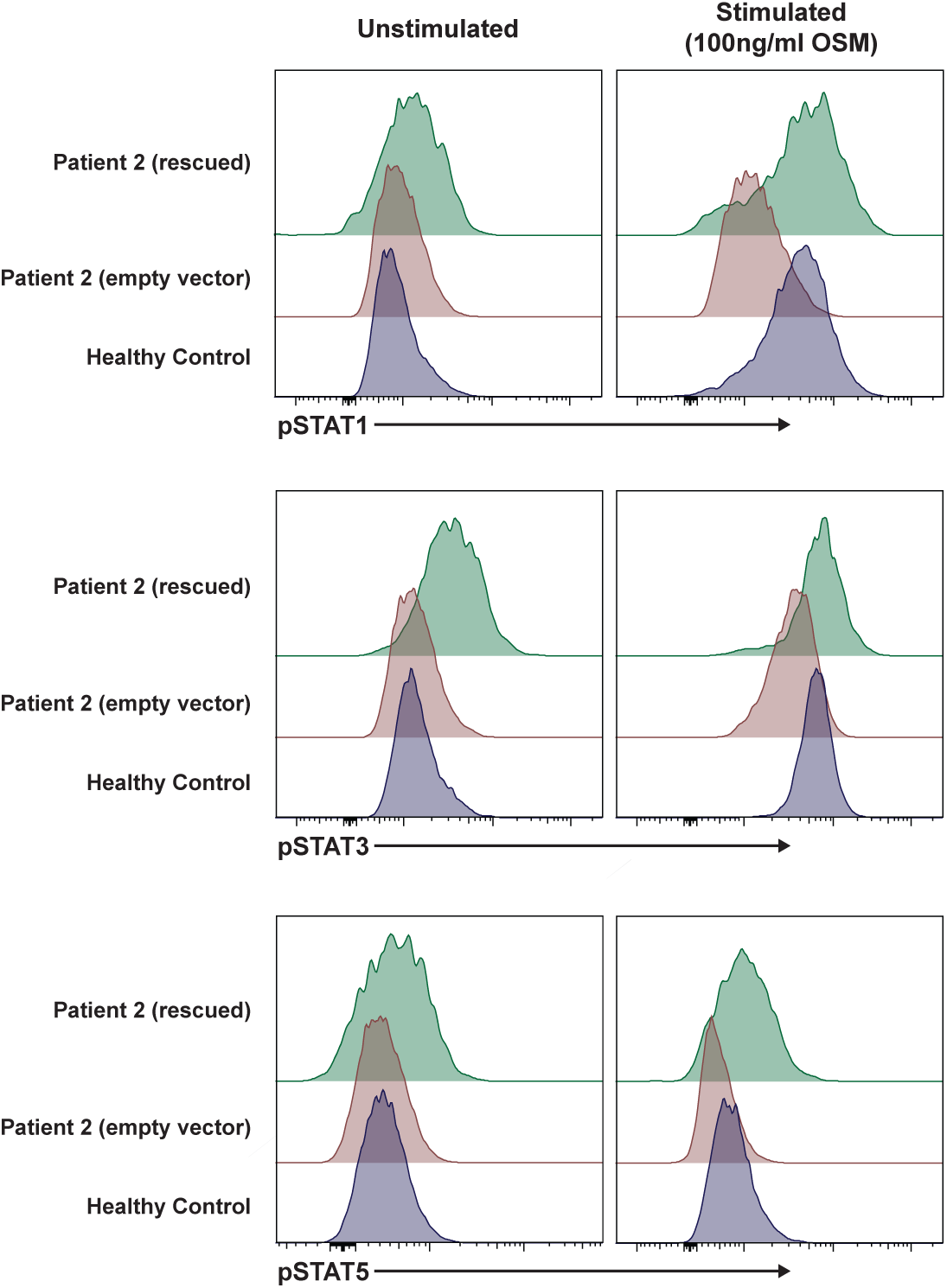
WT-*OSMR* restores STAT signaling in primary fibroblasts. Phosphorylation of STAT1, STAT3, and STAT5 was quantified in primary fibroblasts from Patient 2 that were transduced with WT-*OSMR* or an empty vector and compared to primary fibroblasts from a healthy control before and after treatment with OSM (100ng/ml for 15 minutes).

**Table S1:**
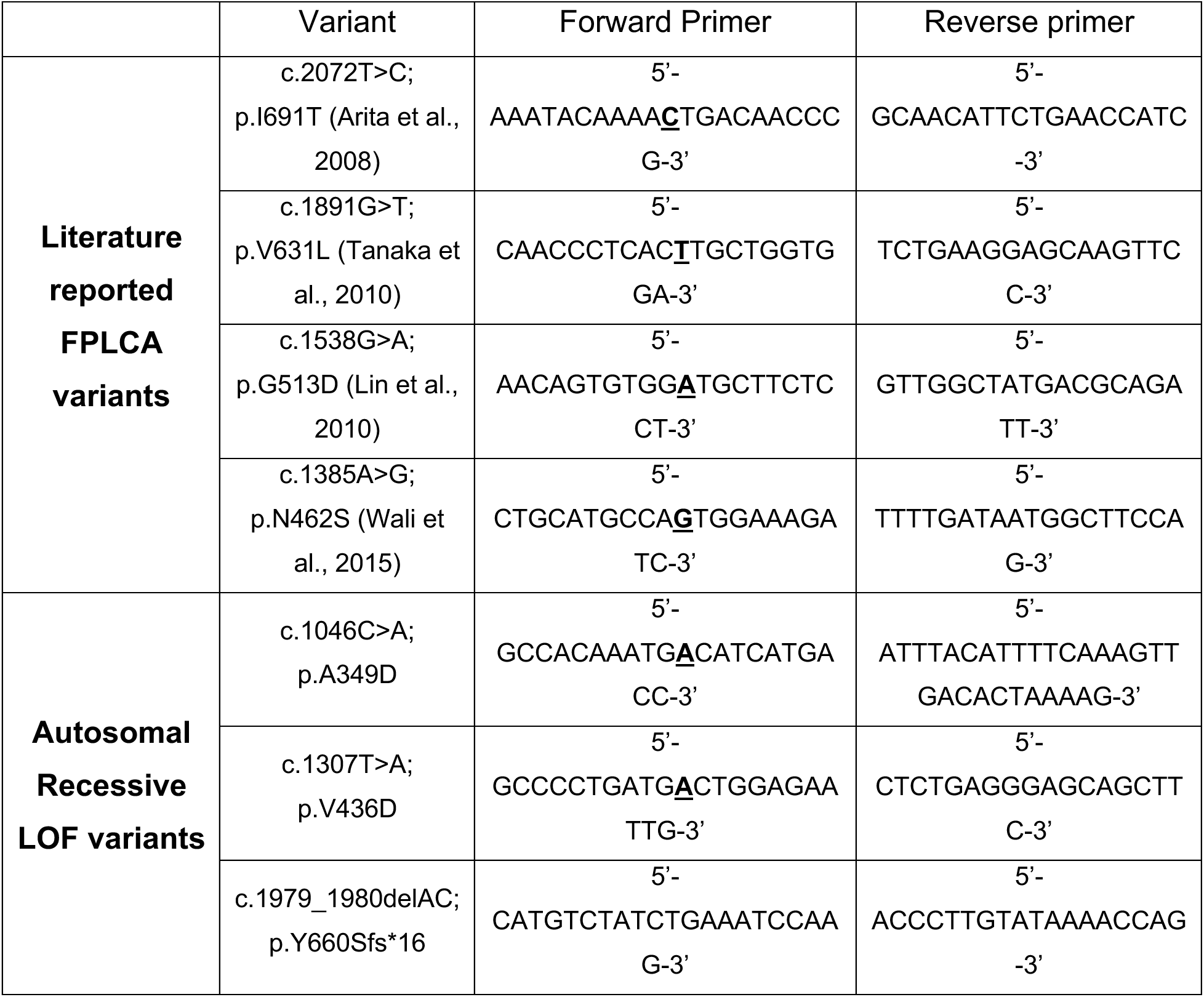
List of primers used for site-directed mutagenesis in the *OSMR* gene. The site of nucleotide change is underlined and bolded in the forward primer sequence.

**Table S2:** List of pathways that are significantly activated in patient and healthy control fibroblasts after stimulation with OSM.

|  | HC stimulated vs<br>Unstimulated |  |  | P2 (EV) stimulated vs<br>Unstimulated |  |  | P2 (Rescued) stimulated<br>vs Unstimulated |  |  |
| --- | --- | --- | --- | --- | --- | --- | --- | --- | --- |
|  | NES | p | q | NES | p | q | NES | p | q |
| **Gamma_IFN** | 2.71 | <0.001 | <0.001 | 2.27 | <0.001 | <0.001 | 2.56 | <0.001 | <0.001 |
| **Alpha_IFN** | 2.61 | <0.001 | <0.001 | 2.13 | <0.001 | <0.001 | 2.52 | <0.001 | <0.001 |
| **JAK-STAT3** | 2.17 | <0.001 | <0.001 | 1.6 | 0.027 | 0.04 | 2.05 | <0.001 | <0.001 |
| **Allo_reject** | 2.15 | <0.001 | <0.001 | 1.53 | 0.014 | 0.059 | 2.07 | <0.001 | <0.001 |
| **Inflam_resp** | 2.11 | <0.001 | <0.001 | 1.45 | 0.028 | 0.092 | 2.11 | <0.001 | <0.001 |
| **TNF_NFkB** | 1.94 | <0.001 | <0.001 | -1.77 | <0.001 | 0.025 | 1.83 | <0.001 | 0.002 |
| **IL2-STAT5** | 1.88 | <0.001 | 0.001 | 1.43 | 0.021 | 0.099 | 1.86 | <0.001 | 0.001 |
| **Complement** | 1.8 | <0.001 | 0.001 | -1.19 | 0.119 | 0.337 | 1.77 | 0.001 | 0.004 |
| **MYC_V2** | 1.73 | 0.003 | 0.005 | 1.06 | 0.369 | 0.625 | 1.46 | 0.052 | 0.089 |
| **G2M** | 1.7 | <0.001 | 0.007 | 1.95 | <0.001 | 0.002 | 0.55 | 1 | 1 |
| **E2F** | 1.64 | <0.001 | 0.015 | 1.71 | <0.001 | 0.018 | -0.98 | 0.57 | 1 |

EV = empty vector; HC = healthy control; NES = normalized enrichment scores.

